# Projected Impact of Concurrently Available Long-Acting Injectable and Daily-Oral HIV Pre-Exposure Prophylaxis: A Mathematical Model

**DOI:** 10.1101/19012443

**Authors:** Kevin M. Maloney, Adrien Le Guillou, Robert A. Driggers, Supriya Sarkar, Emeli J. Anderson, Amyn A. Malik, Samuel M. Jenness

**Author notes:** Deceased (Robert A. Driggers). **CORRESPONDING AUTHOR**: Kevin Maloney, MPH, Alternate: Samuel M. Jenness.

## Abstract

**Background:** Long-acting injectable HIV pre-exposure prophylaxis (LAI-PrEP) is reportedly efficacious, although full trial results have not been published. We used a dynamic network model of HIV transmission among men who have sex with men (MSM) to assess the population impact of LAI-PrEP when available concurrently with daily-oral (DO) PrEP.

**Methods:** The reference model represents the current HIV epidemiology and DO-PrEP coverage (15% among indicated) among MSM in the southeastern US. Primary analyses investigated varied PrEP uptake and proportion selecting LAI-PrEP. Secondary analyses evaluated uncertainty in pharmacokinetic efficacy and LAI-PrEP persistence relative to DO-PrEP.

**Results:** Compared to the reference scenario, if 50% chose LAI-PrEP, 4.3% (95% SI: −7.3%, 14.5%) of infections would be averted over 10 years. LAI-PrEP impact is slightly greater than the DO-PrEP only regimen based on assumptions of higher adherence and partial protection after discontinuation. If the total PrEP initiation rate doubled, 17.1% (95% SI: 6.7%, 26.4%) of infections would be averted. The highest population-level impact occurred when LAI-PrEP uptake and persistence improved.

**Conclusions:** If LAI-PrEP replaces DO-PrEP, its availability will modestly improve the population impact. LAI-PrEP will make a more substantial impact if its availability drives higher total PrEP coverage, or if persistence is greater for LAI-PrEP.

**Summary:** Long-acting injectable pre-exposure prophylaxis (LAI-PrEP) will modestly decrease HIV incidence if it replaces daily-oral PrEP use among men who have sex with men. LAI-PrEP will make a more substantial impact if its availability drives higher total PrEP coverage.

## BACKGROUND

Men who have sex with men (MSM) remain at increased risk for HIV in the United States, accounting for two-thirds of new infections in 2018 [1]. Pre-exposure prophylaxis (PrEP) is a highly effective method to reduce HIV acquisition [2, 3]. However, implementation challenges undermine the potential for PrEP to reduce incidence at the population level. For example, PrEP uptake remains low [4-7]. The US Centers for Disease Control and Prevention (CDC) estimate that 1.1 million US adults were behaviorally indicated to use PrEP in 2017 [8], but only 100,000 accessed PrEP [4]. Efforts to increase PrEP use are undermined by poor persistence, resulting in early discontinuation among individuals with ongoing HIV risk [9-11]. Previous PrEP models have predicted that current coverage levels are insufficient to reach many HIV prevention targets [12, 13]. Among MSM accessing PrEP, inadequate adherence further reduces its population-level prevention benefits [10, 14].

Alternative PrEP products that are more acceptable and easier to use for some may address these implementation challenges. Daily-oral PrEP (DO-PrEP) is currently the only dosing strategy recommended by the CDC and the US Food and Drug Administration [15]. However, alternative dosing regimens [16] have proven efficacious; novel formulations are in various stages of development, including long-acting injectable [17] and topical formulations [18], implants and other devices [19]. Cabotegravir is an investigational antiretroviral (strand-transfer integrase inhibitor) [17]. When formulated as a long-acting injectable suspension (LAI-PrEP), cabotegravir is maintained with periodic dosing at 8-week intervals [17]. The Phase III clinical trial for LAI-PrEP efficacy among MSM (HPTN-083) has been discontinued early with fewer incident infections observed among MSM randomized to LAI-PrEP compared to DO-PrEP [20], although full trial outcomes have yet to be released.

LAI-PrEP may address adherence challenges with DO-PrEP because the pill burden would be eliminated [14, 17]. However, the clinical burden for LAI-PrEP may be greater for some because intramuscular injections are administered by a healthcare professional at 8-week intervals (compared to 12-week intervals for clinical monitoring of DO-PrEP) [15]. LAI-PrEP may be more acceptable for users unwilling or unable to take a daily pill [21-25]. Estimates of preference for LAI-PrEP range from 30% to 67% [21, 23, 25-28], but real-world selection of LAI-PrEP over DO-PrEP is unknown. Inferring population-level benefits from hypothetical LAI-PrEP preferences depends on the interaction of uptake and persistence.

Mathematical modeling provides insights into the mechanisms by which LAI-PrEP may differ from DO-PrEP under these conditions of uncertainty. A recent study [29] simulated the population impact when either DO-PrEP or LAI-PrEP were allocated to MSM. The study found that in scenarios with LAI-PrEP there was a greater decline in incidence than in scenarios with DO-PrEP. However, the impact when LAI-PrEP is available as an additional option to DO-PrEP remains unclear. It was also unclear how challenges to persistence will limit the population impact of LAI-PrEP which, unlike DO-PrEP, provides continued but waning protection even after discontinuation [17]. Since publication of this model, pharmacokinetic data from a Phase II clinical trial of LAI-PrEP has become available [17], which can be used to specify model parameters.

To estimate the potential population-level impact of LAI-PrEP, we used a network-based mathematical model of HIV transmission dynamics among MSM in the southeastern US. To understand how concurrent availability of LAI-PrEP and DO-PrEP could maximize HIV prevention, we simulated scenarios varying overall PrEP uptake and the proportion choosing LAI-PrEP versus DO-PrEP. We also conducted sensitivity analyses of the LAI-PrEP biological parameters (pharmacokinetic and efficacy) and behavioral parameters (discontinuation rate compared to DO-PrEP).

## METHODS

We used a mathematical model of HIV transmission dynamics among black, white, and Hispanic MSM, aged 15-65 years, in the Atlanta area over a 10-year period. The model was simulated using *EpiModel* [30], a software platform for modeling disease dynamics in sexual networks using temporal exponential random graph models (TERGMs) [31]. Building on past PrEP models [12], we developed the model framework and parameterization for LAI-PrEP to explore differences with DO-PrEP. Full methodological details can be found in the Web Appendix.

### Sexual Network Model

All network and behavioral parameters were drawn from the ARTnet study, a 2017-2019 egocentric network study of 4,904 MSM reporting on 16,198 sexual partnerships, including duration, concurrency and other attributes [32]. The model was initialized with 10,000 MSM, but the network size varied based on entry (sexual debut) and exit (mortality or aging out at 65). Summary statistics from ARTnet were fit to statistical models under the TERGM framework to simulate one-off, casual, and main partnerships. Partnership formation occurred stochastically by partnership type, age and race mixing, number of ongoing partnerships, and sorting by receptive and insertive positioning. Relational dissolution was modeled with a constant hazard based on the median duration for partnership type.

For MSM with HIV, rates were assigned for diagnosis, initiation of antiretroviral therapy (ART), and viral suppression [33, 34]. HIV disease progressed through the acute, chronic and AIDS stages. HIV viral loads were modeled continuously, in the absence of ART. ART decreased HIV viral loads, with a corresponding decrease in the probability of HIV transmission [35, 36]. Within serodiscordant sexual partnerships, HIV transmission occurred stochastically with a per-act probability based on PrEP use [14], condom use [37], sexual positioning [38], and circumcision status of the insertive partner [39].

### PrEP Uptake, Adherence, and Persistence

Uptake of PrEP was determined using a two-step process. Parameters for LAI-PrEP and DO-PrEP are shown in **Table 1**. First, we specified the overall probability of initiating PrEP. Second, we specified the probability of choosing LAI-PrEP versus DO-PrEP. By simulating uptake and selection using separate probabilities, we were able to explore counterfactual scenarios regarding how user decisions interact. Eligibility to initiate PrEP of either formulation was based on CDC guidelines for DO-PrEP indications [12, 15]. For MSM using DO-PrEP, we assigned a probability distribution for medication adherence corresponding to low (< 2 doses/week, 8.9%), medium (2–3 doses/week, 12.7%), and high adherence (≥ 4 doses/week, 78.4%) [40], which modified the level of protection against HIV acquisition (hazard ratios: 0.69, 0.19, and 0.02, respectively) [14]. PrEP users were screened for HIV every 90 days according to CDC guidelines [15].

**Table 1.** Pre-exposure prophylaxis parameters in reference model, base LAI-PrEP parameters and LAI-PrEP values varied in counterfactual scenarios.

| Parameter | Reference Scenario | Base LAI-PrEP Scenario | Counterfactual LAI-PrEP Scenarios | Reference |
| --- | --- | --- | --- | --- |
| **Initiation and Persistence** |  |  |  |  |
| Prevalence of overall PrEP use given indication | 15.1% | 15.1% | Varies | [45-47] |
| Probability of initiating PrEP per time-step | 0.411% | 0.411% | 0 – 100% |  |
| Probability of selecting LAI-PrEP | 0% | 50% | 0 – 100% | [21, 23, 25-28] |
| Spontaneous discontinuation rate, median time | 224 days | 224 days | 75 – 672 days | [41] |
| **DO-PrEP** |  |  |  |  |
| Adherence level, % of DO-PrEP users |  |  |  | [40] |
| Low (< 2 doses/week) | 8.9% | --- | --- |  |
| Medium (2-3 doses/week) | 12.7% | --- | --- |  |
| High (4+ doses/week) | 78.4% | --- | --- |  |
| Efficacy, per-exposure hazard-ratio |  |  |  | [14] |
| Low adherence | 0.69 | --- | --- |  |
| Medium adherence | 0.19 | --- | --- |  |
| High adherence | 0.02 | --- | --- |  |
| **LAI-PrEP** |  |  |  |  |
| Peak plasma concentration ($P$) | --- | 3.59 µg/ml | 1.5 – 6.5 µg/ml | [17] |
| Half-life ($T_{1/2}$) | --- | 35 days | 15 – 60 days | [17] |
| Days since last injection ($t$) | --- | Varies | Varies | |
| Plasma concentration ($P_t$) | --- | $P_t = P * \left(\frac{1}{2}\right)^{\frac{t}{T_{1/2}}}$ | --- | [17] |
| Efficacy, per-exposure hazard-ratio | --- | $HR = e^{P_t * B}$ | ± 5% – 50% | |
| Beta ($B$) | --- | -9.06 | --- | [42] |
| Hazard-ratio at 4 x PA-IC<sub>90</sub> | --- | 0.002 | Varies | [42] |

Based on CDC guidelines, behavioral indications were reassessed annually; if indications had lapsed, PrEP would be discontinued for both DO-PrEP and LAI-PrEP [15]. We also allowed spontaneous discontinuation of DO-PrEP to occur based on an observational study showing 57% of PrEP patients remained in care 6 months after initiation [41]. We translated this into a constant probability of discontinuation assuming a geometric distribution to calculate the median time to stoppage (224 days) [41]. It is unknown whether MSM using LAI-PrEP would have the same persistence as has been observed among MSM using DO-PrEP, so our base LAI-PrEP conditions assumed that the rate of spontaneous discontinuation of LAI-PrEP would be the same as the rate assigned to DO-PrEP.

### Injectable PrEP Pharmacokinetics and Efficacy

Based on studies of nonhuman primates, the target cabotegravir plasma concentration in clinical trials was set to 4-times the protein-adjusted 90% inhibitory concentration (PA-IC_90_) [42-44]. A Phase IIa clinical trial showed that 600-mg intramuscular injections at 8-week intervals were sufficient to maintain the target plasma concentration [17]. For simplicity, we assumed that men achieved the target peak plasma concentration following the first injection (in clinical trials, the second injection was administered after 4-weeks to initially achieve the target concentration) and without an initial oral safety phase.

Using data from the Phase Ila trial to model the pharmacokinetics of injectable cabotegravir [17], we estimated the median peak plasma concentration (3.59 μg/ml) following each injection after and including the second injection, as well as the rate of decay in longitudinal follow-up. We estimated a continuous half-life function (35 days) of drug elimination based on these two parameters. This estimate shows that plasma concentrations above PA-IC_90_ and 4-times PA-IC_90_ are maintained for 22 and 12 weeks, respectively, with limited drug still present 52 weeks after the final injection.

The Phase Ill trial results for cabotegravir as LAI-PrEP have not been published. Thus, we used the results of a macaque study to model the relative reduction in the per-exposure probability of HIV infection [42]. We used logistic regression to estimate the probability of infection given the plasma concentration of cabotegravir at the time of exposure. We transformed the results to the human equivalent of the PA-IC_90_ to obtain a single parameter for the relative reduction in infection probability given the plasma concentration of cabotegravir. We estimated that LAI-PrEP is greater than 99% effective at 4-times PA-IC_90_ (HR = 0.002).

### Reference Model and Counterfactual Scenarios

The reference model represents current DO-PrEP uptake, in which approximately 15% of indicated MSM in the southeastern US use PrEP [45-47]. We calibrated the model to maintain a steady state prevalence of DO-PrEP use. To reach 15% DO-PrEP, the probability of initiating PrEP given indications was set to 0.411% per time-step, with LAI-PrEP selection set to 0%.

Our primary analysis explored scenarios of overall PrEP uptake by simultaneously varying the probability of PrEP initiation (supplementing current DO-PrEP uptake by relative factors of 0 to 6 times the base parameter) and the probability of selecting LAI-PrEP (replacing current DO-PrEP uptake from 0% to 100%). The probability of selecting LAI-PrEP was fixed at 50% for all secondary analyses, while the probability of initiating PrEP was fixed at the reference model value. Second, to better understand the pharmacological properties of LAI-PrEP, we varied the pharmacokinetic and efficacy parameters. We varied the LAI-PrEP pharmacokinetics by simultaneously modifying the peak plasma concentration after injection (1.5 to 6.5 μg/ml) and the half-life of drug elimination (15 to 60 days). Separately, we varied the efficacy of LAI-PrEP by scaling the per-exposure hazard ratio for transmission (5%, 10%, 25%, and 50% increase or decrease in efficacy compared to the base parameter). Finally, to understand how the effectiveness of LAI-PrEP could depend on continued use during periods of sexual risk, we varied the rate of spontaneous discontinuation. For this, we modified the median time using LAI-PrEP from 33% (approximately 75 days) to 300% (672 days) and calculated new spontaneous discontinuation rates.

### Calibration, Simulation, and Analysis

The network was initialized with 10,000 MSM and calibrated to the estimated prevalence of diagnosed HIV (33% for black MSM; 27% for Hispanic MSM; and 8.4% for white MSM) [48]. Each model scenario was simulated over 10 years and for 250 times. The outcomes reported were HIV prevalence and incidence per 100 person-years at risk at the end of 10 years, and the percent of infections averted (PIA). The PIA was calculated by comparing the cumulative incidence in each counterfactual scenario to the reference model. We summarized the results using the median values of all simulations and 95% simulation intervals (SI).

## RESULTS

In the reference scenario, 15.1% (95% SI: 14.4%, 15.9%) of indicated MSM were using DO-PrEP (0% LAI-PrEP). At the end of the 10-year simulation, the HIV incidence was 1.19 (95% SI: 0.93, 1.50) per 100 person-years and prevalence was 22.4% (95% SI: 21.5%, 23.4%). Increasing both the probability of initiating PrEP (supplementing current DO-PrEP use) and the probability of selecting LAI-PrEP (replacing current DO-PrEP use) would result in fewer infections over 10 years, compared to the reference scenario (**Figure 1**). The benefit of introducing LAI-PrEP is greater in scenarios where overall PrEP uptake also increased. Small improvements, however, would still be realized at lower proportions of MSM initiating PrEP and selecting LAI-PrEP.

**Figure 1.**
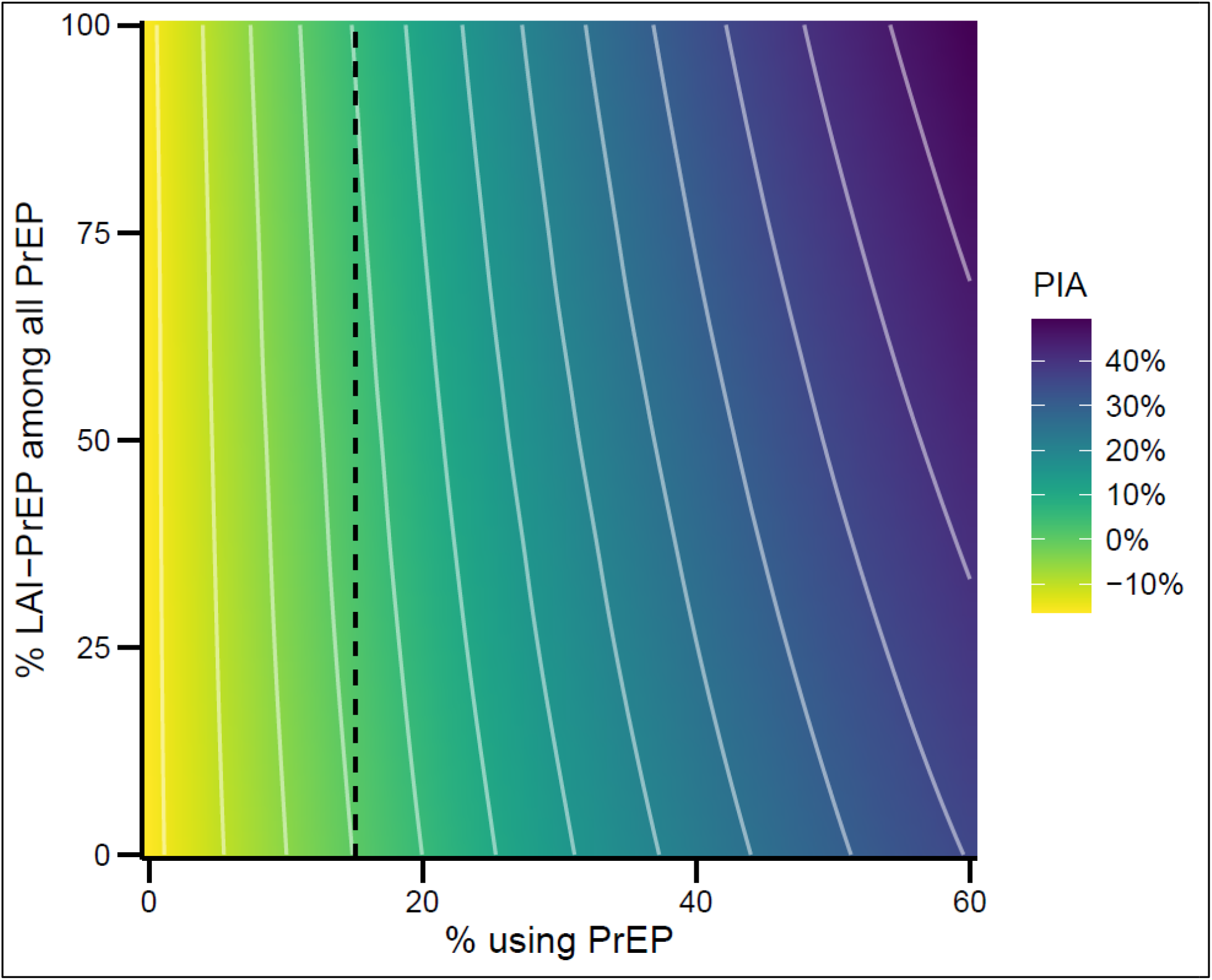
Population benefit of varying overall PrEP uptake and proportion using LAI-PrEP. Legend: Percent of infections averted (PIA) over 10 years, compared to the reference scenario of 15% (dashed line) of behaviorally indicated men who have sex with men using pre-exposure prophylaxis and 0% choosing long-acting injectable pre-exposure prophylaxis (LAI-PrEP) versus daily-oral pre-exposure prophylaxis, across varying overall PrEP uptake and proportion using LAI-PrEP.

Increasing either or both the initiation and the LAI-PrEP selection probabilities will increase the proportion of indicated MSM protected from PrEP, which in turn causes HIV incidence to decline (**Table 2**, with additional scenarios in **Supplemental Table 14**). With PrEP limited to DO-PrEP, increasing the probability of initiating PrEP by two-fold resulted in a greater proportion of indicated MSM using DO-PrEP (26.7%; 95% SI: 25.8%, 27.5%) and 11.3% (95% SI: 0.1%, 21.1%) of infections averted. Alternatively, when the probability of selecting LAI-PrEP was increased to 50% and the PrEP initiation rate was held constant, the proportion of MSM using DO-PrEP dropped to 7.6% (95% SI: 7.1%, 8.2%) while LAI-PrEP use increased to 8.4% (95% SI: 7.9%, 9.0%). In addition to the MSM actively using LAI-PrEP, 7.3% (95% SI: 6.8%, 7.7%) of indicated MSM had waning plasma concentration and thus partial protection after discontinuing LAI-PrEP. This scenario resulted in a modest decrease in incidence (4.3% of infections averted; 95% SI: −7.3%, 14.5%). While the median result indicates an improvement, the lower bound of the 95% SI shows that incidence increased for many of the scenarios with LAI-PrEP, relative to the median value of the reference model; similarly, the upper bound of the interval shows that a larger decrease in incidence is possible.

**Table 2.**
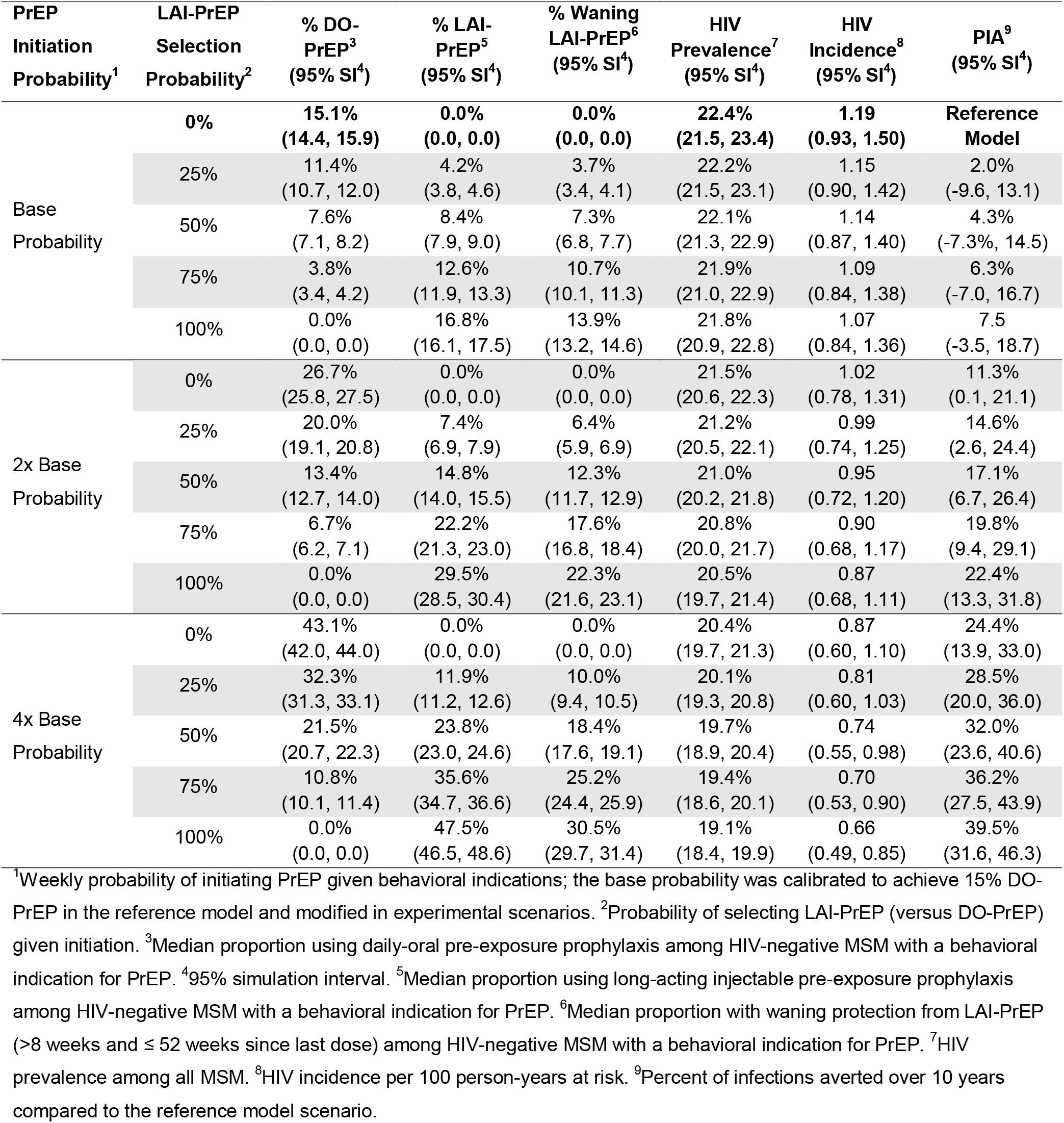
Varying rate of pre-exposure prophylaxis initiation and proportion choosing long-acting injectable pre-exposure prophylaxis (LAI-PrEP) versus daily-oral (DO) PrEP compared to the reference scenario of current uptake of DO-PrEP.

| PrEP Initiation Probability<sup>1</sup> | LAI-PrEP Selection Probability<sup>2</sup> | % DO-PrEP<sup>3</sup><br>(95% SI<sup>4</sup>) | % LAI-PrEP<sup>5</sup><br>(95% SI<sup>4</sup>) | % Waning LAI-PrEP<sup>6</sup><br>(95% SI<sup>4</sup>) | HIV Prevalence<sup>7</sup><br>(95% SI<sup>4</sup>) | HIV Incidence<sup>8</sup><br>(95% SI<sup>4</sup>) | PIA<sup>9</sup><br>(95% SI<sup>4</sup>) |
| --- | --- | --- | --- | --- | --- | --- | --- |
| Base Probability | 0% | 15.1%<br>(14.4, 15.9) | 0.0%<br>(0.0, 0.0) | 0.0%<br>(0.0, 0.0) | 22.4%<br>(21.5, 23.4) | 1.19<br>(0.93, 1.50) | Reference Model |
|  | 25% | 11.4%<br>(10.7, 12.0) | 4.2%<br>(3.8, 4.6) | 3.7%<br>(3.4, 4.1) | 22.2%<br>(21.5, 23.1) | 1.15<br>(0.90, 1.42) | 2.0%<br>(-9.6, 13.1) |
|  | 50% | 7.6%<br>(7.1, 8.2) | 8.4%<br>(7.9, 9.0) | 7.3%<br>(6.8, 7.7) | 22.1%<br>(21.3, 22.9) | 1.14<br>(0.87, 1.40) | 4.3%<br>(-7.3%, 14.5) |
|  | 75% | 3.8%<br>(3.4, 4.2) | 12.6%<br>(11.9, 13.3) | 10.7%<br>(10.1, 11.3) | 21.9%<br>(21.0, 22.9) | 1.09<br>(0.84, 1.38) | 6.3%<br>(-7.0, 16.7) |
|  | 100% | 0.0%<br>(0.0, 0.0) | 16.8%<br>(16.1, 17.5) | 13.9%<br>(13.2, 14.6) | 21.8%<br>(20.9, 22.8) | 1.07<br>(0.84, 1.36) | 7.5<br>(-3.5, 18.7) |
| 2x Base Probability | 0% | 26.7%<br>(25.8, 27.5) | 0.0%<br>(0.0, 0.0) | 0.0%<br>(0.0, 0.0) | 21.5%<br>(20.6, 22.3) | 1.02<br>(0.78, 1.31) | 11.3%<br>(0.1, 21.1) |
|  | 25% | 20.0%<br>(19.1, 20.8) | 7.4%<br>(6.9, 7.9) | 6.4%<br>(5.9, 6.9) | 21.2%<br>(20.5, 22.1) | 0.99<br>(0.74, 1.25) | 14.6%<br>(2.6, 24.4) |
|  | 50% | 13.4%<br>(12.7, 14.0) | 14.8%<br>(14.0, 15.5) | 12.3%<br>(11.7, 12.9) | 21.0%<br>(20.2, 21.8) | 0.95<br>(0.72, 1.20) | 17.1%<br>(6.7, 26.4) |
|  | 75% | 6.7%<br>(6.2, 7.1) | 22.2%<br>(21.3, 23.0) | 17.6%<br>(16.8, 18.4) | 20.8%<br>(20.0, 21.7) | 0.90<br>(0.68, 1.17) | 19.8%<br>(9.4, 29.1) |
|  | 100% | 0.0%<br>(0.0, 0.0) | 29.5%<br>(28.5, 30.4) | 22.3%<br>(21.6, 23.1) | 20.5%<br>(19.7, 21.4) | 0.87<br>(0.68, 1.11) | 22.4%<br>(13.3, 31.8) |
| 4x Base Probability | 0% | 43.1%<br>(42.0, 44.0) | 0.0%<br>(0.0, 0.0) | 0.0%<br>(0.0, 0.0) | 20.4%<br>(19.7, 21.3) | 0.87<br>(0.60, 1.10) | 24.4%<br>(13.9, 33.0) |
|  | 25% | 32.3%<br>(31.3, 33.1) | 11.9%<br>(11.2, 12.6) | 10.0%<br>(9.4, 10.5) | 20.1%<br>(19.3, 20.8) | 0.81<br>(0.60, 1.03) | 28.5%<br>(20.0, 36.0) |
|  | 50% | 21.5%<br>(20.7, 22.3) | 23.8%<br>(23.0, 24.6) | 18.4%<br>(17.6, 19.1) | 19.7%<br>(18.9, 20.4) | 0.74<br>(0.55, 0.98) | 32.0%<br>(23.6, 40.6) |
|  | 75% | 10.8%<br>(10.1, 11.4) | 35.6%<br>(34.7, 36.6) | 25.2%<br>(24.4, 25.9) | 19.4%<br>(18.6, 20.1) | 0.70<br>(0.53, 0.90) | 36.2%<br>(27.5, 43.9) |
|  | 100% | 0.0%<br>(0.0, 0.0) | 47.5%<br>(46.5, 48.6) | 30.5%<br>(29.7, 31.4) | 19.1%<br>(18.4, 19.9) | 0.66<br>(0.49, 0.85) | 39.5%<br>(31.6, 46.3) |

<sup>1</sup>Weekly probability of initiating PrEP given behavioral indications; the base probability was calibrated to achieve 15% DO-PrEP in the reference model and modified in experimental scenarios. <sup>2</sup>Probability of selecting LAI-PrEP (versus DO-PrEP) given initiation. <sup>3</sup>Median proportion using daily-oral pre-exposure prophylaxis among HIV-negative MSM with a behavioral indication for PrEP. <sup>4</sup>95% simulation interval. <sup>5</sup>Median proportion using long-acting injectable pre-exposure prophylaxis among HIV-negative MSM with a behavioral indication for PrEP. <sup>6</sup>Median proportion with waning protection from LAI-PrEP (>8 weeks and ≤ 52 weeks since last dose) among HIV-negative MSM with a behavioral indication for PrEP. <sup>7</sup>HIV prevalence among all MSM. <sup>8</sup>HIV incidence per 100 person-years at risk. <sup>9</sup>Percent of infections averted over 10 years compared to the reference model scenario.

If 100% of the MSM accessing PrEP were to select LAI-PrEP, and initiation remained at the reference level, then 16.8% (95% SI: 16.1%, 17.5%) would have full protection from LAI-PrEP and 13.9% (95% SI: 13.2%, 14.6%) would have waning protection, resulting in 7.5% of infections averted (95% SI: −3.5%, 18.7%). Finally, if the probability of initiating PrEP increased two-fold and the proportion choosing LAI-PrEP was 50%, then incidence would decrease with 17.1% (95% SI: 6.7%, 26.4%) of infections averted.

The underlying estimates for LAI-PrEP pharmacokinetics are sensitive to uncertain parameter values (**Figure 2**, **Table 3**, and **Supplemental Table 15**). If the peak plasma concentration were reduced to 2 μg/ml and the half-life were reduced to 15 days (an extreme scenario, well below the *a priori* clinical trial targets), then the modest effect of introducing LAI-PrEP at 50% selection probability (4.3% of infections averted; 95% SI: −7.3%, 14.5%) would be almost eliminated (1.4% of infections averted; 95% SI: −11.5%, 11.9%). When parameters were modified within a smaller range around our estimates (e.g., 3 μg/ml peak plasma concentration and 25 days half-life) then the impact would be limited (3.1% of infections averted; 95% SI: - 10.4%, 13.5%). Similarly, the population impact of LAI-PrEP is sensitive to our estimate of the drug efficacy (**Figure 3, Table 3**, and **Supplemental Table 16**). For example, a 50% decrease in LAI-PrEP efficacy resulted in increased incidence with −4.4% (95% SI: −17.4%, 8.4%) of infections averted. However, a 10% decrease in efficacy would result in a more modest decline in the population benefit of introducing LAI-PrEP (2.4% of infections averted; 95% SI: −9.7%, 13.8%).

**Figure 2.**
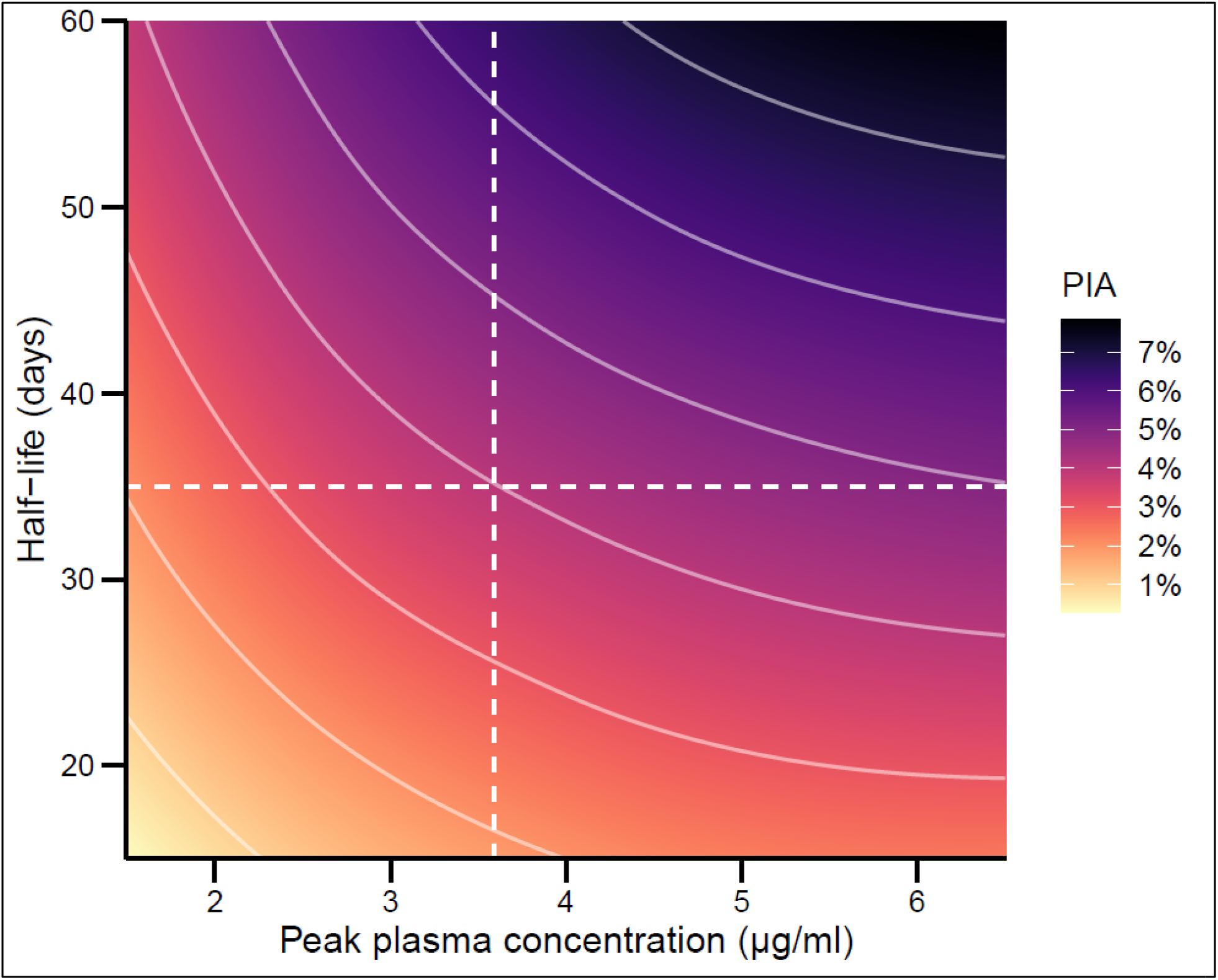
Population benefit of long-acting injectable pre-exposure prophylaxis (LAI-PrEP) across varying pharmacokinetic parameters. Legend: Percent of infections averted (PIA) over 10 years, compared to the reference scenario of 15% (dashed line) of behaviorally indicated men who have sex with men using pre-exposure prophylaxis and 0% choosing long-acting injectable pre-exposure prophylaxis (LAI-PrEP) versus daily-oral pre-exposure prophylaxis, if 50% of PrEP users choose LAI-PrEP when available, across varying parameters for LAI-PrEP pharmacokinetics. Dashed lines indicate peak plasma concentration (3.59 μg/ml) and half-life (5 weeks) used in the base model parameters.

**Figure 3.**
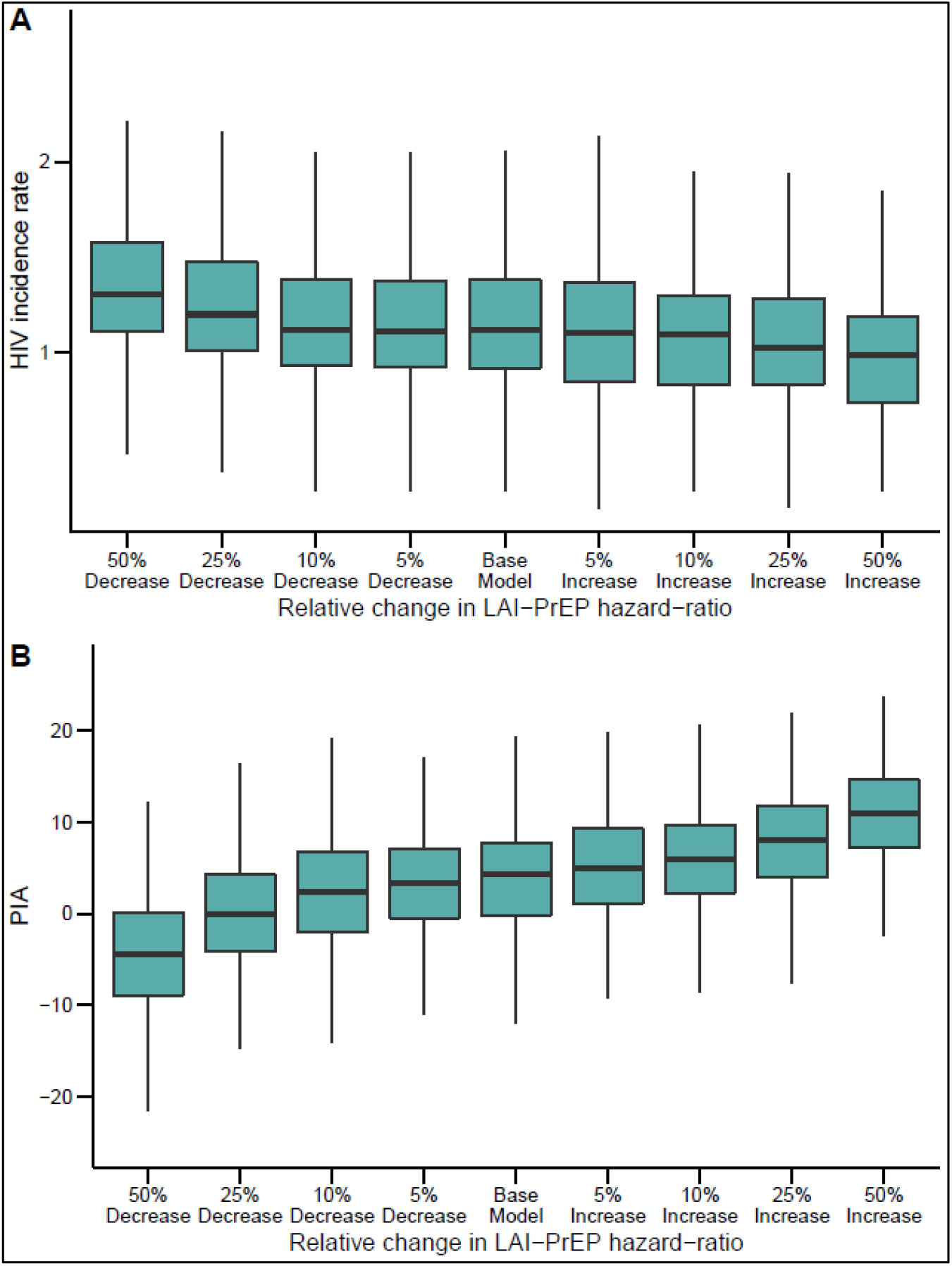
Population benefit of long-acting injectable pre-exposure prophylaxis (LAI-PrEP) across varying efficacy. Legend: HIV outcomes among all men who have sex with men (MSM) in a population with 15% of behaviorally indicated men using PrEP and 50% choosing long-acting injectable pre-exposure prophylaxis (LAI-PrEP) versus daily-oral PrEP, across varying scenarios of the relative change in perexposure LAI-PrEP efficacy. Panel A: HIV incidence rate per 100 person-years at risk. Panel B: Percent of infections averted compared to the reference scenario of 15% of behaviorally indicated MSM using PrEP and 0% choosing LAI-PrEP.

**Table 3.**
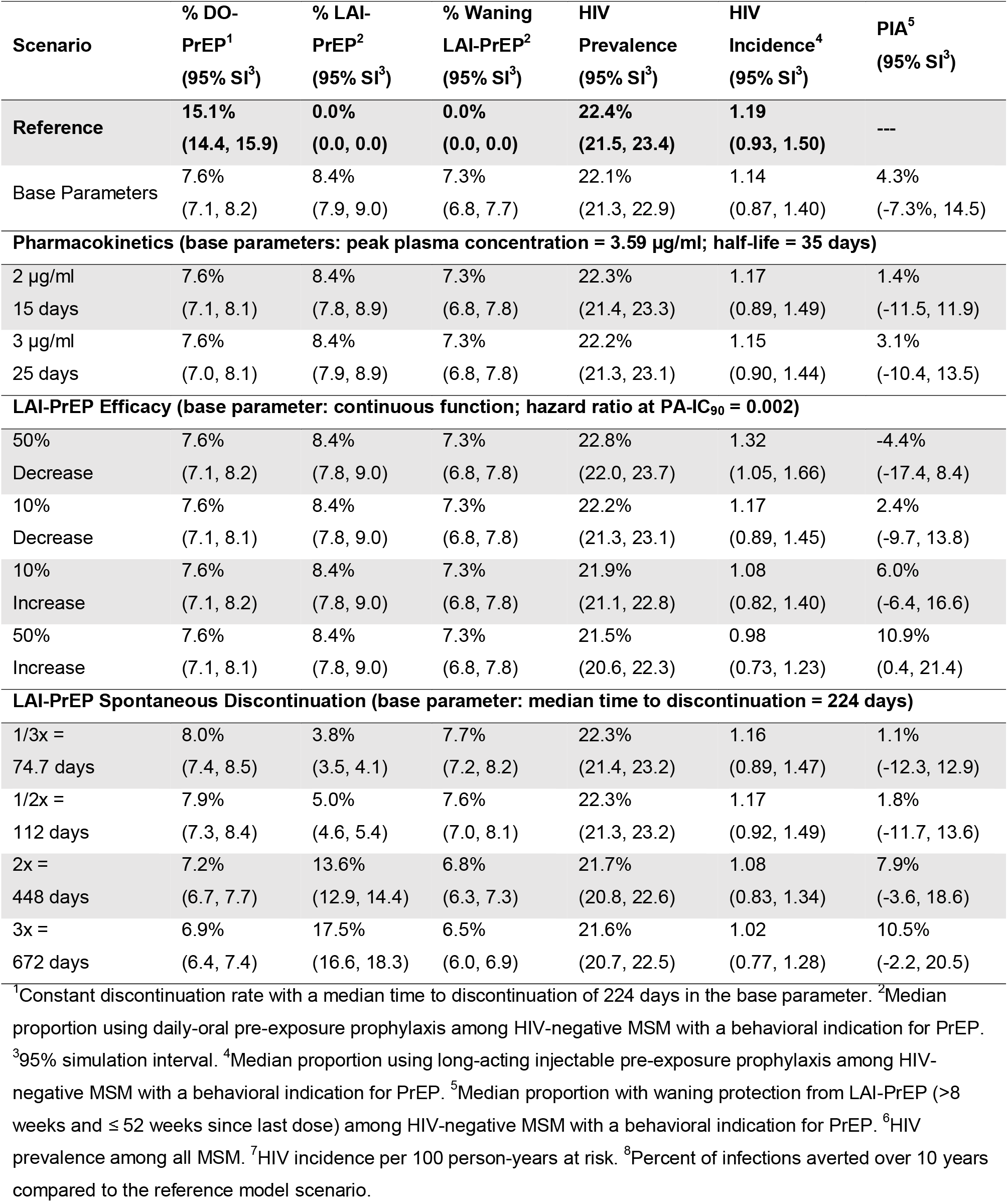
Varying base parameters for long-acting injectable pre-exposure prophylaxis compared to the reference scenario of current uptake of DO-PrEP.

Changes to duration of persistence on LAI-PrEP, relative to the base model duration based on current estimates of DO-PrEP persistence, also impacted population-level impact (**Figure 4, Table 3**, and **Supplementary Table 17**). In scenarios where persistence improved for MSM accessing LAI-PrEP, the prevention benefit of introducing LAI-PrEP increased. For example, if the median time to discontinuation were 448 days (versus 224 days for the base parameter), then active LAI-PrEP use would increase from 8.4% (95% SI: 7.9%, 9.0%) to 13.6% (SI: 12.9%, 14.4%) with a predicted 7.9% (95% SI: −3.0%, 19.6%) of infections averted (compared to 4.3% in the base model; 95% SI: −7.3%, 14.5%). Alternatively, as persistence decreased, the prevalence of LAI-PrEP use among indicated MSM also decreased; the proportion with waning protection from LAI-PrEP increased negligibly because overall PrEP uptake was fixed. When the rate of discontinuation was doubled (median time = 112 days), the predicted prevention benefit declined to 1.8% of infections averted, with the 95% SI (−11.7%, 13.6%) showing that many simulations represented null findings or increased incidence.

**Figure 4.**
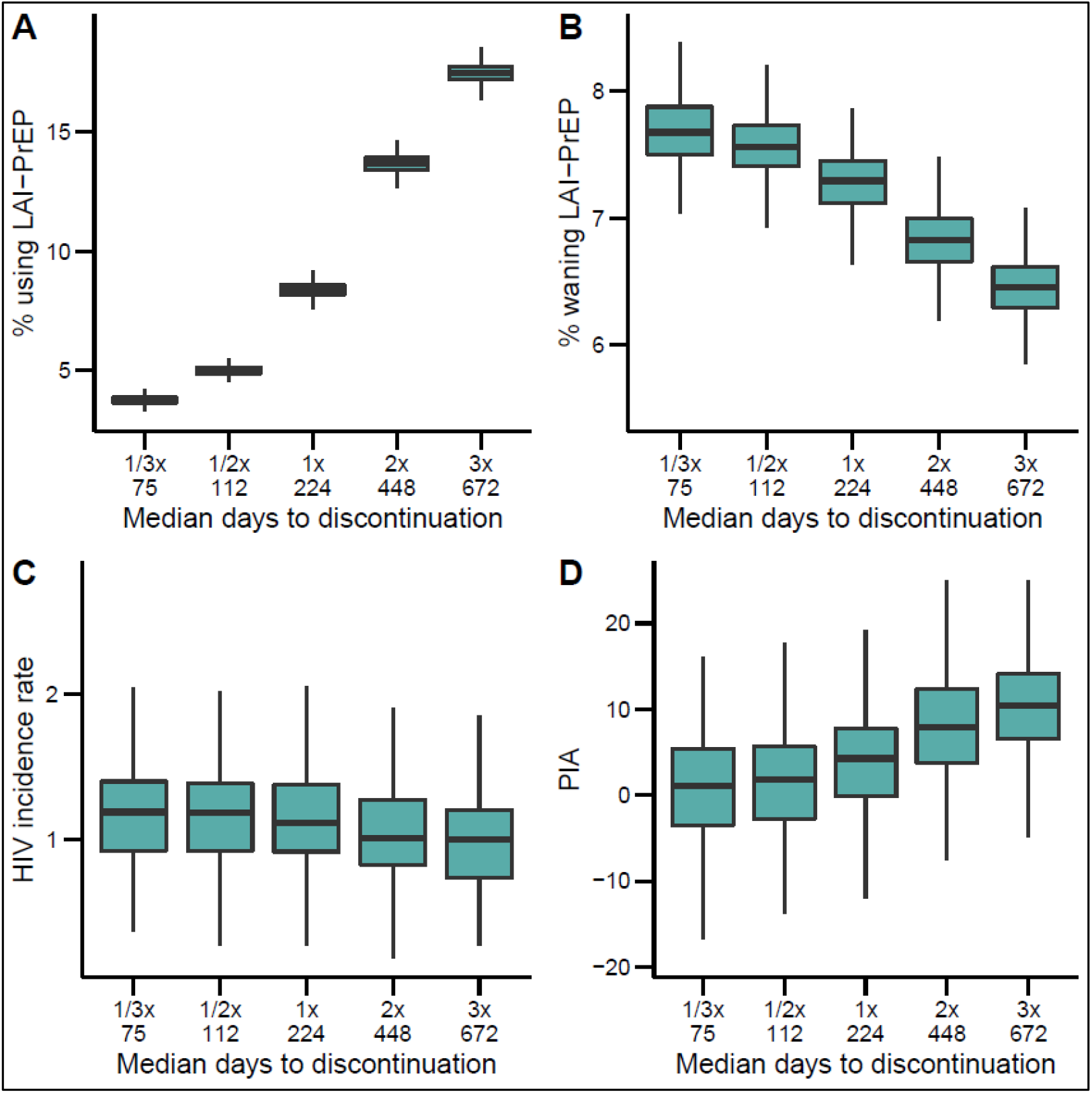
Population benefit of long-acting injectable pre-exposure prophylaxis (LAI-PrEP) across varying rates of spontaneous discontinuation. Legend: Long-acting injectable pre-exposure prophylaxis (LAI-PrEP) use and HIV outcomes among all men who have sex with men (MSM) across scenarios with fixed overall PrEP initiation rate (calibrated to 15% of behaviorally indicated MSM in the reference scenario), 50% choosing LAI-PrEP, and varying LAI-PrEP spontaneous discontinuation rates, compared to a reference scenario of 0% choosing LAI-PrEP. **Panel A**: Percent of behaviorally indicated MSM actively using LAI-PrEP (≤ 8 weeks since last dosage). **Panel B**: Percent of behaviorally indicated MSM with waning protection from LAI-PrEP after discontinuation (> 8 weeks and ≤ 52 weeks since last dosage). **Panel C**: HIV incidence rate per 100 person-years at risk. **Panel D**: Percent of infections averted compared to the reference scenario.

## DISCUSSION

We projected that LAI-PrEP concurrently available with DO-PrEP would modestly reduce HIV incidence among MSM compared to offering only DO-PrEP. The benefit of replacing some DO-PrEP use with LAI-PrEP is due to the elimination of challenges associated with daily adherence to DO-PrEP and the partial protection provided by the long metabolic half-life of LAI-PrEP even after discontinuation. The benefit of LAI-PrEP would increase if uptake supplements current DO-PrEP uptake. To our knowledge, this is the first modeling study to predict the population impact of LAI-PrEP when available concurrently with DO-PrEP.

The introduction of LAI-PrEP could yield small improvements to the prevention benefit of PrEP if uptake replaces current DO-PrEP use, but our model predicts even greater benefit if LAI-PrEP acts to supplement current DO-PrEP uptake. Our results may be generalizable to other PrEP formulations which may replace or supplement existing formulations, although models will need to assess each new formulation as biological and clinical factors may vary. We designed our model to explore PrEP uptake using two components: total PrEP coverage and cross-sectional mix of DO-PrEP and LAI-PrEP. It is unknown how many MSM currently using DO-PrEP will switch to LAI-PrEP once available; however, future preference estimates range from 35–67% would switch to LAI-PrEP [25, 28]. Low uptake has been a limiting factor to the population benefit of DO-PrEP [4-6]. Similar to other studies [12, 13], we found that large-scale reductions in HIV incidence will require greater PrEP uptake among MSM than has currently been observed in the US [4-6]. This may be achieved if the availability of LAI-PrEP increases the proportion of indicated individuals accessing PrEP. Although 66% of PrEP inexperienced MSM indicate interest in LAI-PrEP [27], translating interest to actual uptake has proven difficult for DO-PrEP.

In our model, the prevention benefits of LAI-PrEP were greater when persistence increased above the levels currently observed for DO-PrEP. Our finding is in direct contrast to an earlier LAI-PrEP modeling study, which counterintuitively reported lower HIV incidence as LAI-PrEP persistence decreased [29]. However, that model assumed PrEP was allocated to maintain a constant prevalence of active users in the population so that a new LAI-PrEP user immediately replaced one who had discontinued. As a result, the total person-time on LAI-PrEP would artificially increase in scenarios with lower persistence, through the increased number of individuals with waning protection after discontinuation. This model assumption is unrealistic, and the corresponding relationship between discontinuation and LAI-PrEP impact implausible. In our model, we disentangled these two components of PrEP prevalence (initiation and persistence) in order to evaluate the individual contribution of each factor. We therefore varied LAI-PrEP persistence without artificially inflating the person-time on LAI-PrEP, isolating the effects of initiation and persistence. Our model shows that improving persistence would increase total PrEP coverage in the population and thus decrease HIV incidence, even if the overall PrEP initiation rate remained constant.

Persistence will likely remain a challenge for all persons accessing PrEP. The reasons for discontinuation off DO-PrEP in practice include decreased HIV risk perception, nonadherence to the clinical protocol for PrEP, insurance barriers and experiencing side-effects [9, 49]. The clinical procedures of LAI-PrEP may be even more challenging for some to maintain, given more frequent clinical care visits (i.e., six annual visits for LAI-PrEP compared to four for DO-PrEP). However, the history of contraceptive medication could serve as a guide: increased choices improved uptake, persistence, and adherence, as individuals selected the modality which most closely aligned with their preferences [50]. As options increase for PrEP, uptake, persistence and adherence may similarly improve across the board. When LAI-PrEP becomes available, clinical practice guidelines should be developed to counsel potential users on the formulation that best meets their needs.

### Limitations

Our model has a number of important limitations. The first is uncertainty of the LAI-PrEP parameters, since Phase III clinical trials have not been published. Our model parameters may be misspecified which would cause our primary results to be biased. To accommodate this, we tested a range of plausible parameters for LAI-PrEP pharmacokinetics and efficacy, which had only a small effect on the model results. Our model was also a simplification of the protocol for LAI-PrEP used in the Phase III clinical trial [17]. Increased complexity and clinical burden of the protocol may result in lower persistence. Specifically, we did not include the 4-week oral dosing phase of cabotegravir that is used to ensure safety. We also assumed that the peak plasma concentration for cabotegravir would be achieved after a single intramuscular dose, instead of the two initial doses at a 4-week interval. Similarly, we did not model an oral ramp-down phase for individuals discontinuing LAI-PrEP under the supervision of their clinician (opposed to spontaneous discontinuation). This latter phase of LAI-PrEP is used to prevent infection during the waning tail of LAI-PrEP protection. Finally, we did not model resistance to cabotegravir among individuals that acquire HIV infection following discontinuation of LAI-PrEP, while the drug remains present but provides only partial protection. This phenomenon presents a new challenge compared to DO-PrEP. Future studies will need to evaluate resistance and what impact this may have on transmission dynamics, if clinical trial and observational data demonstrate the occurrence of resistance.

### Conclusions

The introduction of LAI-PrEP will modestly improve the prevention benefit of DO-PrEP if LAI-PrEP only serves as a replacement to DO-PrEP. This is due to the non-inferiority in the biological benefit of LAI-PrEP over DO-PrEP. However, LAI-PrEP could make a more substantial impact on HIV incidence if the availability of LAI-PrEP increases the likelihood that indicated users will start on PrEP, or decreases the likelihood that current PrEP users will discontinue despite ongoing risk. Prior to the FDA approval of LAI-PrEP, public health policy and implementation science on LAI-PrEP should anticipate these user behaviors as critical to maximizing the benefits of this new HIV prevention tool.

## Data Availability

This work was a simulation of disease transmission and does not represent analysis of primary data. The simulation parameters were derived from a variety of published data sources and referenced throughout the manuscript as necessary.

https://github.com/EpiModel/injectable-prep

## ACKNOWLEDGEMENTS

We would like to acknowledge our friend and colleague, Robert A. Driggers, who contributed substantially to the design and conduct of this study, and in authoring this paper, prior to his unexpected death. Rob was an important member of our team and he will be missed.

## DISCLOSURES

S.S. is an employee of ViiV Healthcare, which owns the patent for Cabotegravir. Her scientific contribution to this study was completed before she joined ViiV Healthcare. ViiV Healthcare did not fund or participate in any stage of this study. K.M., R.D., E.A., A.M., and S.J. have no conflicts to report.

## AUTHORSHIP

K.M., R.D., S.S., E.A., and A.M. conceived the analysis and assisted in coding the model. K.M. conducted the statistical analyses and drafted the manuscript. R.D. provided critical input with K.M. to specify the pharmacokinetic properties of the LAI-PrEP model. A.L.G. assisted in coding the model and provided input to the analysis plan. S.J. provided direction to the development of the analysis plan, led the coding of the model, and provided critical input to the manuscript and technical appendix. All authors reviewed and provided substantial feedback to drafts of the manuscript.

## FUNDING

This work was supported by National Institutes of Health grants R21 MH112449, R01 AI138783, and P30 AI050409 (Emory University Center for AIDS Research).

## CORRESPONDING AUTHOR

Kevin Maloney, MPH Department of Epidemiology Emory University 1518 Clifton Rd. Atlanta, GA 30322 Phone: (404) 727-0483 Fax: (404) 712-8392

## MEETINGS

A previous version of this model was presented as a poster at the International AIDS Society’s 10th Conference on HIV Science (Abstract ID: TUPEC390). A pre-print of this manuscript has been posted to medRxiv (medrxiv.org/content/10.1101/19012443v1).

## SUPPLEMENTAL DIGITAL CONTACT

Maloney_Projected_Impact_of_Injectable_PrEP_Appendix_version_1.docx

## Notes

### Summary of Updates

Reviewer suggestions/comments

## REFERENCES

1. Centers for Disease Control and Prevention. Estimated HIV incidence and prevalence in the United States, 2014-2018. HIV Surveillance Supplemental Report. Vol. 25, 2020.

2. Grant RM, Lama JR, Anderson PL, et al. Preexposure chemoprophylaxis for HIV prevention in men who have sex with men. N Engl J Med 2010; 363:2587–99.

3. Spinner CD, Brunetta J, Shalit P, et al. DISCOVER study for HIV pre-exposure prophylaxis (PrEP): F/TAF has a more rapid onset and longer sustained duration of HIV protection compared with F/TDF. 10th IAS Conference on HIV Science. Mexico City, Mexico, 2019.

4. Sullivan PS, Giler RM, Mouhanna F, et al. Trends in the use of oral emtricitabine/tenofovir disoproxil fumarate for pre-exposure prophylaxis against HIV infection, United States, 2012-2017. Ann Epidemiol 2018; 28:833–40.

5. Finlayson T, Cha S, Xia M, et al. Changes in HIV Preexposure Prophylaxis Awareness and Use Among Men Who Have Sex with Men - 20 Urban Areas, 2014 and 2017. MMWR Morb Mortal Wkly Rep 2019; 68:597–603.

6. Kamitani E, Wichser ME, Adegbite AH, et al. Increasing prevalence of self-reported HIV preexposure prophylaxis use in published surveys: a systematic review and meta-analysis. AIDS 2018; 32:2633–5.

7. Parsons JT, Rendina HJ, Lassiter JM, Whitfield TH, Starks TJ, Grov C. Uptake of HIV PreExposure Prophylaxis (PrEP) in a National Cohort of Gay and Bisexual Men in the United States. J Acquir Immune Defic Syndr 2017; 74:285–92.

8. Smith DK, Van Handel M, Grey J. Estimates of adults with indications for HIV pre-exposure prophylaxis by jurisdiction, transmission risk group, and race/ethnicity, United States, 2015. Ann Epidemiol 2018; 28:850–7 e9.

9. Krakower D, Maloney KM, Powell VE, et al. Patterns and clinical consequences of discontinuing HIV preexposure prophylaxis during primary care. J Int AIDS Soc 2019; 22:e25250.

10. Marcus JL, Hurley LB, Hare CB, et al. Preexposure Prophylaxis for HIV Prevention in a Large Integrated Health Care System: Adherence, Renal Safety, and Discontinuation. J Acquir Immune Defic Syndr 2016; 73:540–6.

11. Marcus JL, Hurley LB, Nguyen DP, Silverberg MJ, Volk JE. Redefining Human Immunodeficiency Virus (HIV) Preexposure Prophylaxis Failures. Clin Infect Dis 2017; 65:1768–9.

12. Jenness SM, Goodreau SM, Rosenberg E, et al. Impact of the Centers for Disease Control's HIV Preexposure Prophylaxis Guidelines for Men Who Have Sex With Men in the United States. J Infect Dis 2016; 214:1800–7.

13. Sullivan PS, Carballo-Dieguez A, Coates T, et al. Successes and challenges of HIV prevention in men who have sex with men. Lancet 2012; 380:388–99.

14. Grant RM, Anderson PL, McMahan V, et al. Uptake of pre-exposure prophylaxis, sexual practices, and HIV incidence in men and transgender women who have sex with men: a cohort study. Lancet Infect Dis 2014; 14:820–9.

15. Centers for Disease Control and Prevention. Preexposure prophylaxis for the prevention of HIV infection in the United States - 2017 update: a clinical practice guideline, 2018.

16. Molina JM, Capitant C, Spire B, et al. On-Demand Preexposure Prophylaxis in Men at High Risk for HIV-1 Infection. N Engl J Med 2015; 373:2237–46.

17. Landovitz RJ, Li S, Grinsztejn B, et al. Safety, tolerability, and pharmacokinetics of long-acting injectable cabotegravir in low-risk HIV-uninfected individuals: HPTN 077, a phase 2a randomized controlled trial. PLoS Med 2018; 15:e1002690.

18. Baeten J. Topical and on-demand PrEP. 10th IAS Conference on HIV Science. Mexico City, Mexico: International AIDS Society, 2019.

19. Flexner CW. Implants and transdermal drug delivery systems for HIV prevention. 10th IAS Conference on HIV Science. Mexico City, Mexico: International AIDS Society, 2019.

20. Landovitz RJ. HPTN083 interim results: Pre-exposure prophylaxis (PrEP) containing long-acting injectable cabotegravir (CAB-LA) is safe and highly effective for cisgender men and transgender women who have sex with men (MSM,TGW). 23rd International AIDS Conference. Virtual, 2020.

21. Parsons JT, Rendina HJ, Whitfield TH, Grov C. Familiarity with and Preferences for Oral and Long-Acting Injectable HIV Pre-exposure Prophylaxis (PrEP) in a National Sample of Gay and Bisexual Men in the U.S. AIDS Behav 2016; 20:1390–9.

22. Biello K, Coffey-Esquivel J, Hosek S, et al. Development of a mobile-based application to increase uptake of HIV testing among young US men who have sex with men. J Int Aids Soc 2016; 19.

23. Greene GJ, Swann G, Fought AJ, et al. Preferences for Long-Acting Pre-exposure Prophylaxis (PrEP), Daily Oral PrEP, or Condoms for HIV Prevention Among U.S. Men Who Have Sex with Men. AIDS Behav 2017; 21:1336–49.

24. Biello KB, Hosek S, Drucker MT, et al. Preferences for Injectable PrEP Among Young U.S. Cisgender Men and Transgender Women and Men Who Have Sex with Men. Arch Sex Behav 2018; 47:2101–7.

25. Meyers K, Wu Y, Brill A, Sandfort T, Golub SA. To switch or not to switch: Intentions to switch to injectable PrEP among gay and bisexual men with at least twelve months oral PrEP experience. PLoS One 2018; 13:e0200296.

26. Levy ME, Patrick R, Gamble J, et al. Willingness of community-recruited men who have sex with men in Washington, DC to use long-acting injectable HIV pre-exposure prophylaxis. PLoS One 2017; 12:e0183521.

27. Biello KB, Mimiaga MJ, Santostefano CM, Novak DS, Mayer KH. MSM at Highest Risk for HIV Acquisition Express Greatest Interest and Preference for Injectable Antiretroviral PrEP Compared to Daily, Oral Medication. AIDS Behav 2018; 22:1158–64.

28. John SA, Whitfield THF, Rendina HJ, Parsons JT, Grov C. Will Gay and Bisexual Men Taking Oral Pre-exposure Prophylaxis (PrEP) Switch to Long-Acting Injectable PrEP Should It Become Available? AIDS Behav 2018; 22:1184–9.

29. Marshall BDL, Goedel WC, King MRF, et al. Potential effectiveness of long-acting injectable pre-exposure prophylaxis for HIV prevention in men who have sex with men: a modelling study. Lancet HIV 2018; 5:e498-e505.

30. Jenness SM, Goodreau SM, Morris M. EpiModel: An R Package for Mathematical Modeling of Infectious Disease over Networks. J Stat Softw 2018; 84.

31. Krivitsky PN, Handcock MS. A Separable Model for Dynamic Networks. J R Stat Soc Series B Stat Methodol 2014; 76:29–46.

32. Weiss KM, Goodreau SM, Morris M, et al. Egocentric sexual networks of men who have sex with men in the United States: Results from the ARTnet study. Epidemics 2020; 30:100386.

33. Beer L, Mattson CL, Bradley H, Shouse RL, Medical Monitoring P. Trends in ART Prescription and Viral Suppression Among HIV-Positive Young Adults in Care in the United States, 2009-2013. J Acquir Immune Defic Syndr 2017; 76:e1-e6.

34. Beer L, Oster AM, Mattson CL, Skarbinski J, Medical Monitoring P. Disparities in HIV transmission risk among HIV-infected black and white men who have sex with men, United States, 2009. AIDS 2014; 28:105–14.

35. Hughes JP, Baeten JM, Lingappa JR, et al. Determinants of per-coital-act HIV-1 infectivity among African HIV-1-serodiscordant couples. J Infect Dis 2012; 205:358–65.

36. Cohen MS, Chen YQ, McCauley M, et al. Prevention of HIV-1 infection with early antiretroviral therapy. N Engl J Med 2011; 365:493–505.

37. Smith DK, Herbst JH, Zhang X, Rose CE. Condom effectiveness for HIV prevention by consistency of use among men who have sex with men in the United States. J Acquir Immune Defic Syndr 2015; 68:337–44.

38. Goodreau SM, Carnegie NB, Vittinghoff E, et al. What drives the US and Peruvian HIV epidemics in men who have sex with men (MSM)? PLoS One 2012; 7:e50522.

39. Wiysonge CS, Kongnyuy EJ, Shey M, et al. Male circumcision for prevention of homosexual acquisition of HIV in men. Cochrane Database Syst Rev 2011:CD007496.

40. Liu AY, Cohen SE, Vittinghoff E, et al. Preexposure Prophylaxis for HIV Infection Integrated With Municipal- and Community-Based Sexual Health Services. JAMA Intern Med 2016; 176:75–84.

41. Chan PA, Mena L, Patel R, et al. Retention in care outcomes for HIV pre-exposure prophylaxis implementation programmes among men who have sex with men in three US cities. J Int AIDS Soc 2016; 19:20903.

42. Andrews CD, Spreen WR, Mohri H, et al. Long-acting integrase inhibitor protects macaques from intrarectal simian/human immunodeficiency virus. Science 2014; 343:1151–4.

43. Andrews CD, Yueh YL, Spreen WR, et al. A long-acting integrase inhibitor protects female macaques from repeated high-dose intravaginal SHIV challenge. Sci Transl Med 2015; 7:270ra4.

44. Radzio J, Spreen W, Yueh YL, et al. The long-acting integrase inhibitor GSK744 protects macaques from repeated intravaginal SHIV challenge. Sci Transl Med 2015; 7:270ra5.

45. Kelley CF, Kahle E, Siegler A, et al. Applying a PrEP Continuum of Care for Men Who Have Sex With Men in Atlanta, Georgia. Clin Infect Dis 2015; 61:1590–7.

46. Goedel WC, Halkitis PN, Greene RE, Hickson DA, Duncan DT. HIV Risk Behaviors, Perceptions, and Testing and Preexposure Prophylaxis (PrEP) Awareness/Use in Grindr-Using Men Who Have Sex With Men in Atlanta, Georgia. J Assoc Nurses AIDS Care 2016; 27:133–42.

47. Harris NS, Johnson AS, Huang YA, et al. Vital Signs: Status of Human Immunodeficiency Virus Testing, Viral Suppression, and HIV Preexposure Prophylaxis - United States, 2013-2018. MMWR Morb Mortal Wkly Rep 2019; 68:1117–23.

48. Rosenberg ES, Purcell DW, Grey JA, Hankin-Wei A, Hall E, Sullivan PS. Rates of prevalent and new HIV diagnoses by race and ethnicity among men who have sex with men, U.S. states, 2013-2014. Ann Epidemiol 2018; 28:865–73.

49. Lankowski AJ, Bien-Gund CH, Patel VV, Felsen UR, Silvera R, Blackstock OJ. PrEP in the Real World: Predictors of 6-Month Retention in a Diverse Urban Cohort. AIDS Behav 2019; 23:1797–802.

50. Gray AL, Smit JA, Manzini N, Beksinska M. Systematic review of contraceptive medicines: “Does choice make a difference?”. WHO RHRU, 2006.

